# Diagnostic performance of attenuated total reflection Fourier-transform infrared spectroscopy for detecting COVID-19 from routine nasopharyngeal swab samples

**DOI:** 10.1101/2021.11.29.21266906

**Authors:** Helinä Heino, Lassi Rieppo, Tuija Männistö, Mikko J. Sillanpää, Vesa Mäntynen, Simo Saarakkala

**Affiliations:** Research Unit of Medical Imaging, Physics and Technology, University of Oulu, Oulu, Finland; Northern Finland Laboratory Centre NordLab, NordLab Oulu, Oulu, Finland; Research Unit of Mathematical Sciences, University of Oulu, Oulu, Finland; Department of Diagnostic Radiology, Oulu University Hospital, Oulu, Finland

**Keywords:** SARS-COV-2, infrared spectroscopy, machine learning, partial least squares discriminant analysis, classification

## Abstract

Severe acute respiratory syndrome coronavirus 2 (SARS-CoV-2) causing global COVID-19 pandemic since 2019 has led to increasing amount of research to study how to do fast screening and diagnosis to efficiently detect COVID-19 positive cases, and how to prevent spreading of the virus. Our research objective was to study whether SARS-CoV-2 could be detected from routine nasopharyngeal swab samples by using attenuated total reflection Fourier-transform infrared (ATR-FTIR) spectroscopy coupled with partial least squares discriminant analysis (PLS-DA). The advantage of ATR-FTIR is that measurements can be conducted without any sample preparation and no reagents are needed. Our study included 558 positive and 558 negative samples collected from Northern Finland. Overall, we found moderate diagnostic performance for ATR-FTIR when polymerase chain reaction (PCR) was used as the gold standard: the average area under the receiver operating characteristics curve (AUROC) was 0.67-0.68 (min. 0.65, max. 0.69) with 20, 10 and 5 k-fold cross validations. Mean accuracy, sensitivity and specificity was 0.62-0.63 (min. 0.60, max. 0.65), 0.61 (min. 0.58, max. 0.65) and 0.64 (min. 0.59, max. 0.67) with 20, 10 and 5 k-fold cross validations. As a conclusion, our study with relatively large sample set clearly indicate that measured ATR-FTIR spectrum contains specific information for SARS-CoV-2 infection (P<0.001 in label permutation test). However, the diagnostic performance of ATR-FTIR remained only moderate, potentially due to low concentration of viral particles in the transport medium. Further studies are needed before ATR-FTIR can be recommended for fast screening of SARS-CoV-2 from routine nasopharyngeal swab samples.

**Importance:** Attenuated total reflection Fourier-transform infrared (ATR-FTIR) spectroscopy coupled with machine learning-based analysis was applied to detect severe acute respiratory syndrome coronavirus 2 (SARS-CoV-2) from nasopharyngeal swab samples originally collected and processed for polymerase chain reaction (PCR) analysis. Even though our results showed moderate performance, we think that our carefully designed and conducted work is valuable in the field of SARS-CoV-2 diagnostics as there were as many as 1116 nasopharyngeal swab samples (558 negative and 558 positive) collected from individual patients in a real clinical setting. The Real clinical setting refers to the fact that the nasopharyngeal swab samples were collected from people with symptoms typical for COVID-19 or asymptomatic individuals exposed to SARS-CoV-2. The presented technique could be relatively easy to use for point-of-care testing, as ATR-FTIR can be performed with a portable machine without sample preparation and machine learning-based model could give a result immediately after ATR-FTIR measurement.

## Introduction

The Global COVID-19 pandemic has raised a desperate need for an accurate, fast and cheap test to efficiently detect infected people to prevent the spreading of coronavirus (1,2). Polymerase chain reaction (PCR) method is the gold standard for detecting severe acute respiratory syndrome coronavirus 2 (SARS-CoV-2) from respiratory secretions (3). However, PCR is not the best modality for quick screening, as it requires certain sample preparation and transportation to centralized laboratories. Therefore, new cost-effective tests for SARS-CoV-2 are being developed. Furthermore, in the future, the possibility to adjust cost-effective test to the novel viruses could provide a way to avoid new infectious diseases developing to a pandemic state.

According to the scientific literature, attenuated total reflection Fourier-transform infrared (ATR-FTIR) spectroscopy is a potentially suitable method for the fast detection of SARS-CoV-2 infection (4–6). In ATR-FTIR measurement, infrared (IR) light is guided to a sample to measure how the sample molecules interact with the IR light. Collected data shows molecular bond vibrations related to the sample chemical composition, i.e., revealing the chemical fingerprint for the studied sample. Besides fast measurement time in ATR-FTIR method (a few minutes), several ATR-FTIR equipments are already portable, which could allow analysis of the biological samples directly in the public places like border control stations, shopping centres and airports.

ATR-FTIR has been used earlier in diverse studies to detect different conditions from human liquid biopsies, such as breast cancer from saliva (7), dengue fever from blood and serum (8) and hepatitis B and C from sera (9). During the last year, ATR-FTIR method has already been applied to detect SARS-CoV-2 from blood (both in serum and plasma (10,11)) and saliva samples (12,13). Even though results from these studies are extremely promising, the number of investigated samples is typically relatively small. For example, in the study by Barauna et al. excellent results were obtained for detecting SARS-CoV-2 infection directly from the pharyngeal swab samples (accuracy, sensitivity, and specificity of 90%, 95%, and 89%, respectively) within a total of 181 samples (12). Furthermore, in the study by Nogueira et al., authors had 65 nasopharyngeal swab samples in viral transport medium 1 (sensitivity 84%, specificity 66% and accuracy 76.9%) and 178 nasopharyngeal swab samples in viral transport medium 2 (sensitivity 87%, specificity 64% and accuracy 78.4%) (14). Despite these promising preliminary results, more research is needed, especially with larger sample size, in order to judge the true diagnostic performance of the ATR-FTIR method in realistic clinical scenario.

Our aim was to investigate the diagnostic performance of the ATR-FTIR spectroscopy to detect the SARS-CoV-2 infection from the same routine nasopharyngeal swab samples that are used in PCR. Compared to earlier studies, our analyzed sample set contained 558 positive and 558 negative samples, making it the largest ATR-FTIR study so far to detect the SARS-CoV-2 from the very same samples that were originally collected and processed for the PCR.

## Materials and methods

### Nasopharyngeal swab samples and ATR-FTIR measurement

The studied nasopharyngeal swab samples were originally collected by Finnish public healthcare in the Northern Ostrobothnia region, Northern Finland, for conducting PCR tests to detect patients with SARS-CoV-2 infection. Residues from these nasopharyngeal swab samples were stored in a freezer (−20 degrees) and thawed prior to the ATR-FTIR measurements. A total of 558 negative and 558 positive samples were included in this study from January 1st 2020 to November 24th 2020. The ethical permissions were obtained both locally from the Ethical Committee of North Ostrobothnia’s Hospital District as well as nationally from the Finnish Medicines Agency (www.fimea.fi).

The PCR analysis, used as the gold standard for the ATR-FTIR measurements, was conducted by the Northern Finland Laboratory Centre NordLab (www.nordlab.fi), Oulu, Finland. The nasopharyngeal swab samples were collected with a flocked swab and immediately immersed in viral transport medium. During the sample collection period patients were instructed to be tested for infection with SARS-CoV-2, if they had any symptoms indicative of COVID-19. Asymptomatic individuals were tested in Finland upon discretion of physicians working in disease control. The samples were then transported in room temperature to the laboratory performing the analyses. The laboratory had no information on patients’ symptoms. The PCR methods were performed according to instructions. Before the PCR measurements, the swab samples were dissolved into the viral transport medium, and the residues from these samples were stored in the freezer for ATR-FTIR measurements. Before ATR-FTIR all the swab samples were inactivated with viral lysis buffer liquid. Bruker Alpha II FTIR spectrometer (Bruker Optics GmbH, Ettlingen, Germany) equipped with an ATR module (Platinum ATR, Bruker Optics GmbH, Ettlingen, Germany) was used to perform ATR-FTIR measurement one-by-one for the every sample after thawing. The spectral resolution was set to 2 cm^-1^, the number of scans to 64, and the collected spectral range was 4000 to 400 cm^-1^. A droplet (volume: 1 µl) of the sample solution was pipetted onto the ATR crystal (Figure 1). A repeat measurement was started immediately after the sample droplet was placed onto the ATR crystal to collect a total of 10 successive spectra. The spectrum from the last measurement was selected into the final data analysis to ensure that the water has evaporated. This measurement procedure was repeated three times for each sample to minimize differences due to pipetting process. The ATR-crystal was cleaned with ethanol and Virkon disinfectant after every measurement procedure when the measurement was completed to prevent sample contamination.

**Figure 1.**
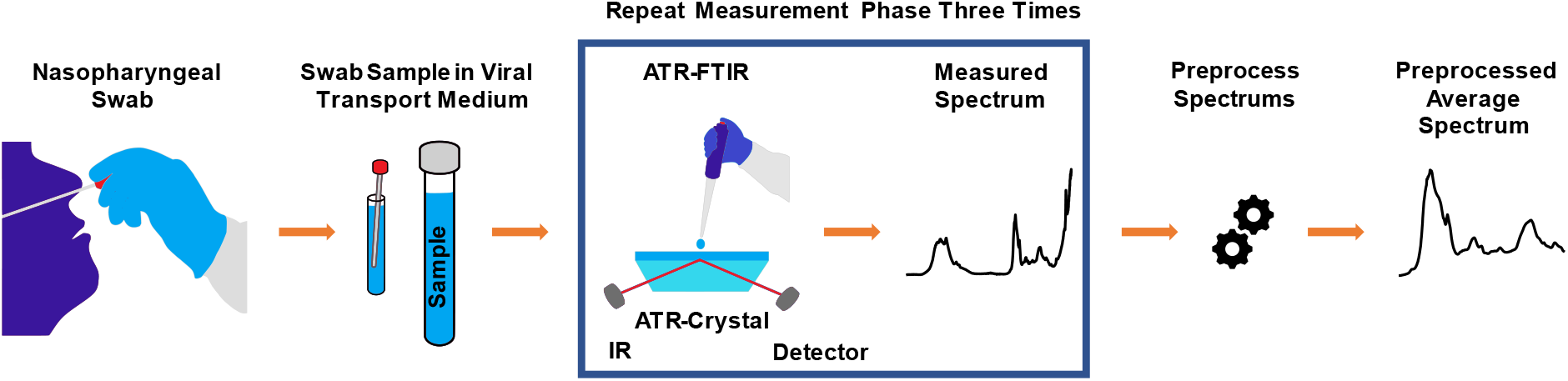
A routine nasopharyngeal swab sample was dissolved into the viral transport medium, analyzed with PCR, and the remnant from that sample was stored in the freezer. The frozen swab sample was thawed and inactivated by adding a viral lysis buffer liquid before the ATR-FTIR measurements. The measurement was conducted by pipetting a one drop from sample solution containing the nasopharyngeal swab sample, the viral lysis buffer and the viral transport medium onto the ATR crystal. Pipetting and measurement was repeated three times to obtain three spectra from each sample. Those three measured spectra were finally averaged so that there was one representative spectrum from each nasopharyngeal swab sample. Before ATR-FTIR measurement, the sample was vortexed to have as homogeneous material distribution as possible, and spun with centrifuge to set the whole sample to the bottom of the tube.

### Spectral preprocessing

The ATR-FTIR spectra were truncated to the fingerprint region of 1800-900 cm^-1^, vector normalized, and the three spectra of each sample were averaged to create one average spectrum per sample. In addition, the spectral region of 1490-1180 cm^-1^ was also analyzed in the same way as fingerprint spectrum region, as preliminary testing indicated similar performance in classification, as with fingerprint spectral region. Preprocessing steps were performed with Anaconda3 (Conda 4.8.3 package manager and Python version 3.8.3) using NumPy, scikit-learn and SciPy packages.

### PLS-DA and data analysis

Partial least squares discriminant analysis (PLS-DA) is a popular method for classification for multivariate datasets. PLS-DA performs dimensionality reduction by creating latent variables, i.e. PLS components, by maximizing the covariance between the new predictor and response scores (15–17). According to authors’ own experience and literature (9,10) PLS-DA is particularly suitable for classification of FTIR spectra, and it was therefore selected as the primary data analysis approach in this study.

Anaconda3 (Conda 4.8.3 package manager and Python version 3.8.3) and PLS Regression package of the scikit-learn library was used to create the PLS-DA model. The performance of the model was studied by using cross-validation with k-fold values 5, 10 and 20. Every k-fold cross-validation was repeated 100 times with different random seed initializations to make sure that the results are not affected by the random seed choice used to divide data into training and validation sets. The presented results have been calculated as average, minimum, and maximum values of the repeated k-fold cross validations. Receiver operating characteristic (ROC) curve, area under the receiver operating characteristics (AUROC) curve, accuracy, sensitivity, specificity, precision, and confusion matrix were used to assess the model performance (18,19).

We repeated the same predictive analysis 1000 times by randomly permuting sample labels in each k-fold case. This permutation analysis was performed to obtain an empirical null distribution of the AUROC values, where we can then see how the extremal position (quantile), the corresponding AUROC value, obtained with the real data, can find in this distribution. This was done to show that the PLS-DA model have the real skill to differentiate between the positive and negative sample groups, and thus it is not possible to obtain a similar magnitude of AUROC values by chance by randomly dividing samples into the negative and positive classes. P-values were calculated by using test 1 according to Ojala and Garriga (20). For permutation tests, see also Efron and Hastie (21). In practice, one random seed used to divide data into the training and validation sets was chosen, and the 1000 permutations were performed to achieve the null distribution of AUROC-values for each k-fold training in question, meaning k-fold 20, 10 or 5. (21)

## Results

The dataset containing ATR-FTIR spectra from the 558 positive and 558 negative nasopharyngeal swab samples, dissolved into the viral transport medium, and inactivated by adding the viral lysis buffer liquid, was analyzed with the PLS-DA method, with PCR analysis as the gold standard method. In the spectral data, the averaged spectra (the mean spectrum from three repetitive measurements per one sample) from the fingerprint region (Figure 2) fed for the PLS-DA model yielded the best results. When the best PLS-DA model performance was obtained, there were 32 latent variables set for the model in case of k-folds 20 and 10, but for k-fold 5 the 31 latent variables were optimal choice (figure 3). The mean AUROC values of 0.68 (min 0.67, max 0.69), 0.68 (min 0.66, max 0.69) and 0.67 (min 0.65, max 0.69) were achieved from the PLS-DA k-fold cross-validation trainings repeated 100 times for k-folds 20, 10 and 5, respectively (Table 1). Obtained P-value for each k-fold showed significant difference between the mean AUROC value achieved in our study, and the mean AUROC value obtained from the same study repeated with the randomly permuted sample labels (for each P-value, P<0.001, see Table 2 from supplementary material). Averaged ROC curves were also calculated (figure 4).

**Table 1.**
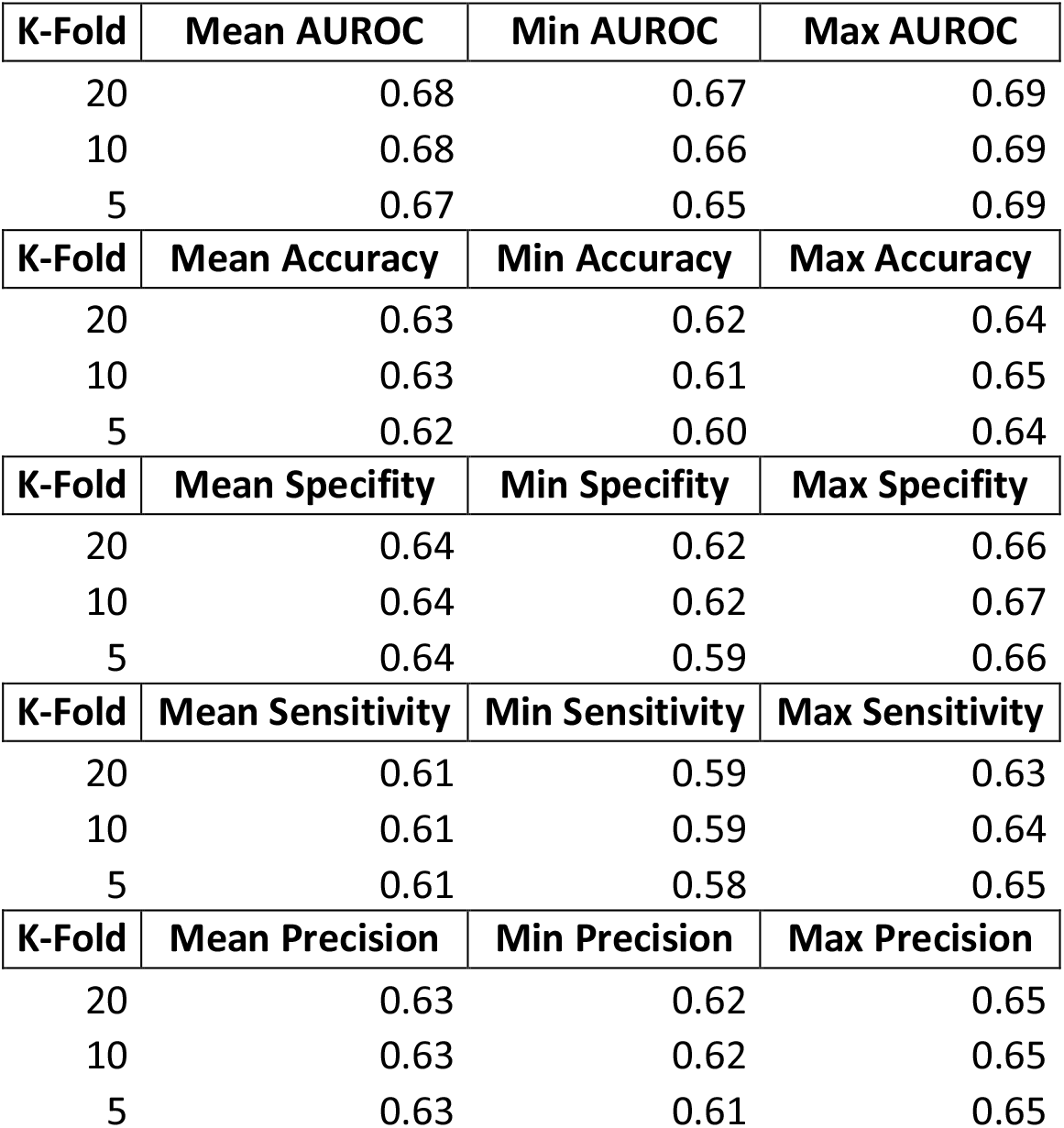
Averaged results from the PLS-DA model k-fold cross-validation predictive ability repeated 100 times with k-fold values 20, 10 and 5. The number of latent variables were set to 32 for k-folds 20 and 10, but for k-fold 5 the latent variables were set to 31, as it was yielding the best performance in predictive ability. There were 558 negative and 558 positive nasopharyngeal swab samples (dissolved into the viral transport medium and inactivated by adding viral lysis buffer liquid) measured with ATR-FTIR spectroscopy. Every sample was measured three times. The measured samples were preprocessed before PLS-DA, by applying spectrum truncation to the spectrum region of 1800-900 cm^-1^, vector normalization and spectrum averaging to create a one average spectrum for every measured nasopharyngeal swab sample.

| K-Fold | Mean AUROC | Min AUROC | Max AUROC |
| --- | --- | --- | --- |
| 20 | 0.68 | 0.67 | 0.69 |
| 10 | 0.68 | 0.66 | 0.69 |
| 5 | 0.67 | 0.65 | 0.69 |
| K-Fold | Mean Accuracy | Min Accuracy | Max Accuracy |
| 20 | 0.63 | 0.62 | 0.64 |
| 10 | 0.63 | 0.61 | 0.65 |
| 5 | 0.62 | 0.60 | 0.64 |
| K-Fold | Mean Specificity | Min Specificity | Max Specificity |
| 20 | 0.64 | 0.62 | 0.66 |
| 10 | 0.64 | 0.62 | 0.67 |
| 5 | 0.64 | 0.59 | 0.66 |
| K-Fold | Mean Sensitivity | Min Sensitivity | Max Sensitivity |
| 20 | 0.61 | 0.59 | 0.63 |
| 10 | 0.61 | 0.59 | 0.64 |
| 5 | 0.61 | 0.58 | 0.65 |
| K-Fold | Mean Precision | Min Precision | Max Precision |
| 20 | 0.63 | 0.62 | 0.65 |
| 10 | 0.63 | 0.62 | 0.65 |
| 5 | 0.63 | 0.61 | 0.65 |

**Table 2.**
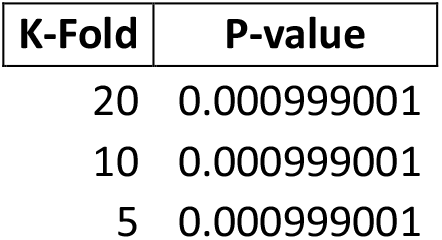
P-values of randomly permuted sample labels for presented mean AUROC values from the fingerprint region. Sample labels were permuted 1000 times when calculating P-values for k-folds 20, 10 and 5. P-values were calculated by using test 1 according to Ojala and Garriga (20).

**Figure 2.**
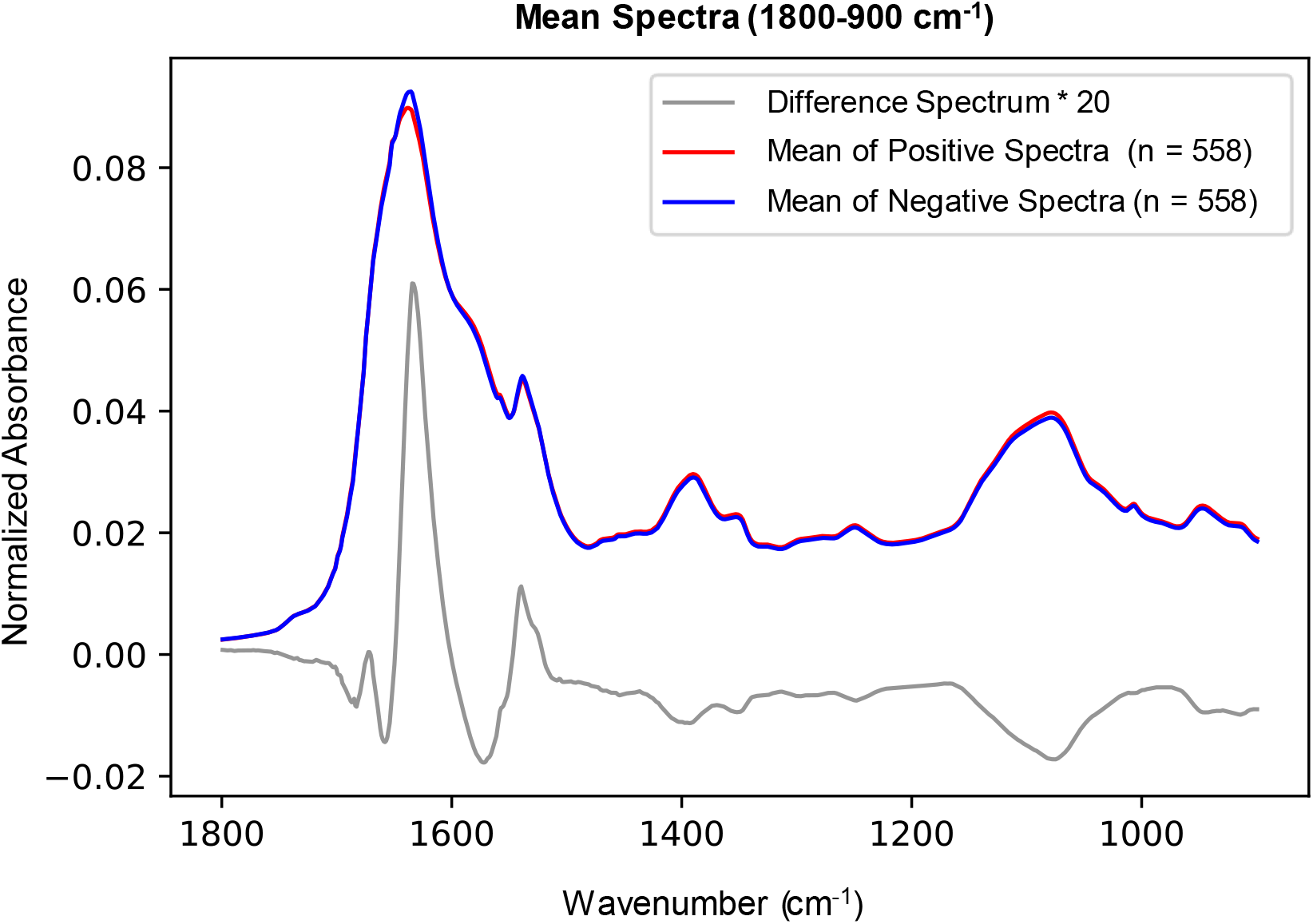
Mean spectra of positive and negative groups from the fingerprint region (1800-900 cm^-1^). Vector normalization was applied before calculating the mean spectra. Difference spectrum multiplied by factor of 20 is also presented to illustrate differences between the sample groups in fingerprint region. Only minor visual differences can be seen in the spectra between the sample groups.

**Figure 3.**
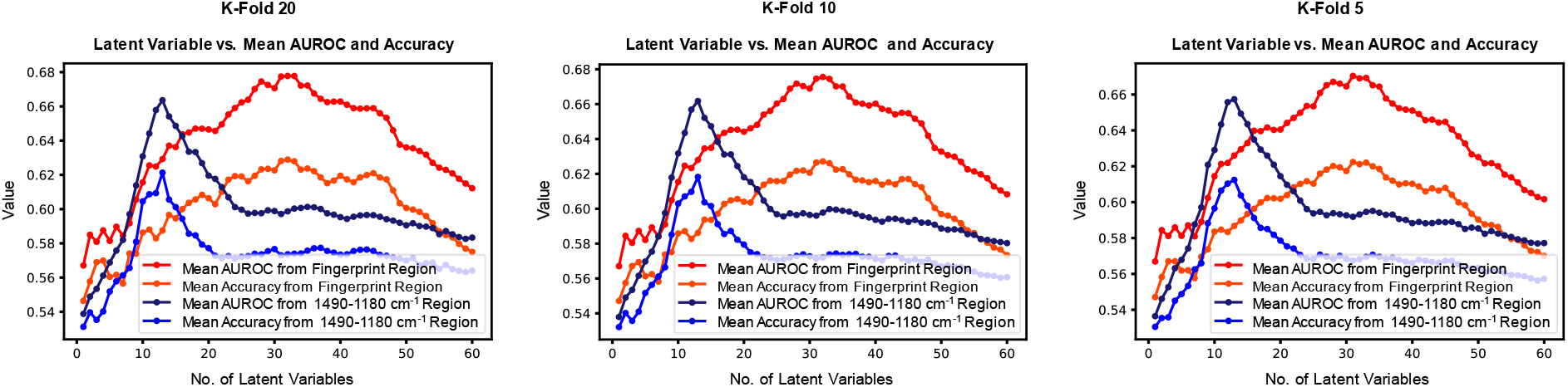
Mean AUROC and mean accuracy values against the latent variables when fingerprint region (1800-900 cm^-1^) and region of 1490-1180 cm^-1^ were studied. As seen from the figure, 32 latent variables yielded the best performance when fingerprint region was studied, and k-fold was 20 or 10, but for k-fold 5 the 31 latent variables were the best possible choice. In case of region of 1490-1180 cm^-1^, the 13 latent variables showed the best performance for k-folds 20, 10 and 5.

**Figure 4.**
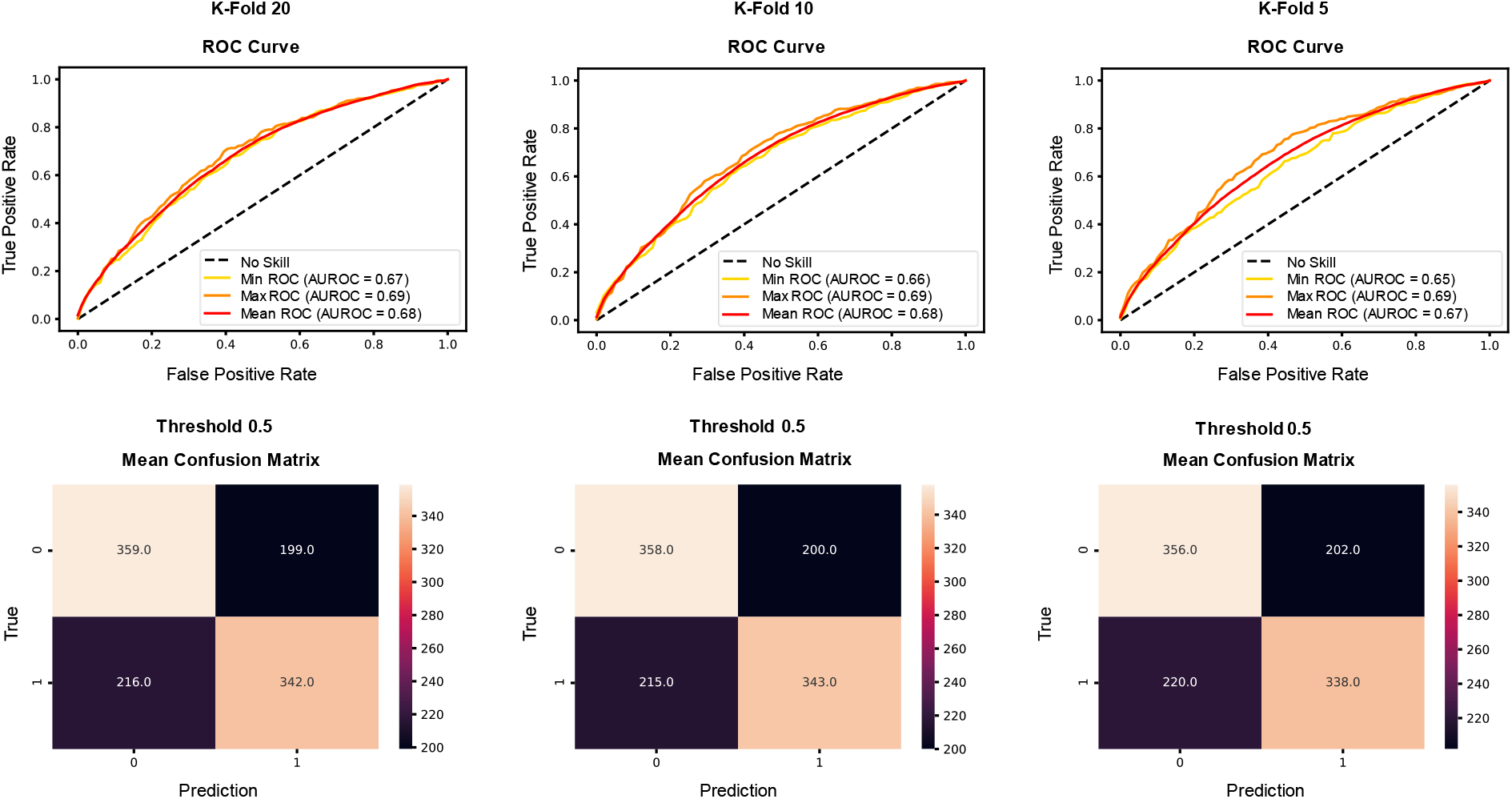
Averaged ROC curves and confusion matrices from PLS-DA k-fold cross-validation predictive ability repeated 100 times with k-folds 20, 10 and 5. Training was accomplished with ATR-FTIR data from the spectral region of 1800-900 cm^-1^.

When the mean accuracies, the mean specificities, the mean sensitivities, and the mean precisions were calculated, a global threshold of 0.5 was set to divide the PLS-DA analyzed samples into the positive and negative classes, also resulting the mean confusion matrixes shown in Figure 4. The mean accuracies of 0.63, 0.63, 0.62, and the mean specificities of 0.64, 0.64 and 0.64 for k-folds 20, 10 and 5, respectively were achieved (Table 1). The mean sensitivities of 0.61, 0.61, 0.61, and the mean precisions of 0.63, 0.63 and 0.63 were obtained for k-folds 20, 10 and 5, respectively (Table 1).

The spectral region of 1490-1180 cm^-1^ (Figure 5) from fingerprint region yielded results close to, or almost the same compared to the fingerprint region when the 13 latent variables were set for the PLS-DA model (figure 3). The achieved mean AUROC values from the PLS-DA training were 0.66 (min 0.66, max 0.67), 0.66 (min 0.65, max 0.67) and 0.66 (min 0.63, max 0.67) for k-folds 20, 10 and 5, respectively (Table 3 in supplementary material). Obtained P-value for each k-fold was showing significant difference between the mean AUROC value from the study of the spectral region of 1490-1180 cm^-1^, and from the same study repeated with randomly permuted sample labels (for each P-value, P<0.001, see Table 4 from supplementary material).

**Table 3.**
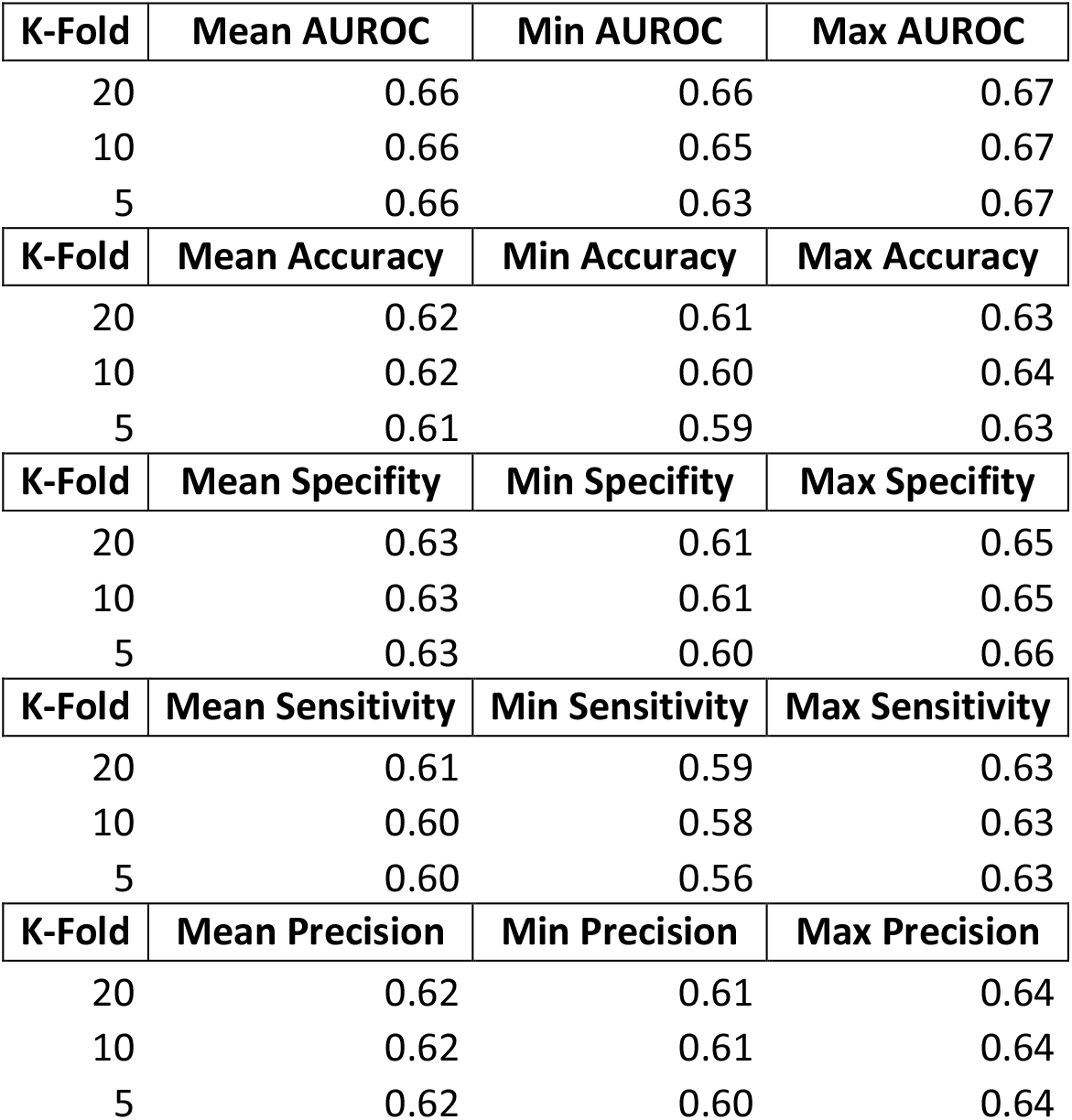
Averaged results from the PLS-DA model k-fold cross-validation predictive ability repeated 100 times with k-fold values 20, 10 and 5. The number of latent variables were set to 13, as it was yielding the best performance in predictive ability. There were 558 negative and 558 positive nasopharyngeal swab samples (dissolved into the viral transport medium and inactivated by adding viral lysis buffer liquid) measured with ATR-FTIR spectroscopy. Every sample was measured three times. The measured samples were preprocessed before PLS-DA, by applying spectrum truncation to the spectrum region of 1490-1180 cm^-1^, vector normalization and spectrum averaging to create a one average spectrum for every measured nasopharyngeal swab sample.

| K-Fold | Mean AUROC | Min AUROC | Max AUROC |
| --- | --- | --- | --- |
| 20 | 0.66 | 0.66 | 0.67 |
| 10 | 0.66 | 0.65 | 0.67 |
| 5 | 0.66 | 0.63 | 0.67 |
| K-Fold | Mean Accuracy | Min Accuracy | Max Accuracy |
| 20 | 0.62 | 0.61 | 0.63 |
| 10 | 0.62 | 0.60 | 0.64 |
| 5 | 0.61 | 0.59 | 0.63 |
| K-Fold | Mean Specificity | Min Specificity | Max Specificity |
| 20 | 0.63 | 0.61 | 0.65 |
| 10 | 0.63 | 0.61 | 0.65 |
| 5 | 0.63 | 0.60 | 0.66 |
| K-Fold | Mean Sensitivity | Min Sensitivity | Max Sensitivity |
| 20 | 0.61 | 0.59 | 0.63 |
| 10 | 0.60 | 0.58 | 0.63 |
| 5 | 0.60 | 0.56 | 0.63 |
| K-Fold | Mean Precision | Min Precision | Max Precision |
| 20 | 0.62 | 0.61 | 0.64 |
| 10 | 0.62 | 0.61 | 0.64 |
| 5 | 0.62 | 0.60 | 0.64 |

**Table 4.**
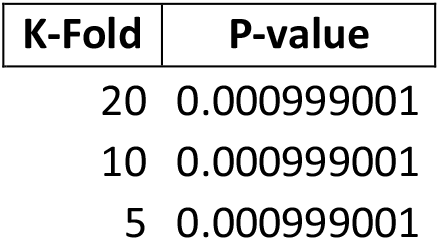
P-values of randomly permuted sample labels for presented mean AUROC values from the spectral region of 1490-1180 cm^-1^. Sample labels were permuted 1000 times when calculating P-values in case of k-folds 20, 10 and 5. P-values were calculated by using test 1 according to Ojala and Garriga (20).

**Figure 5.**
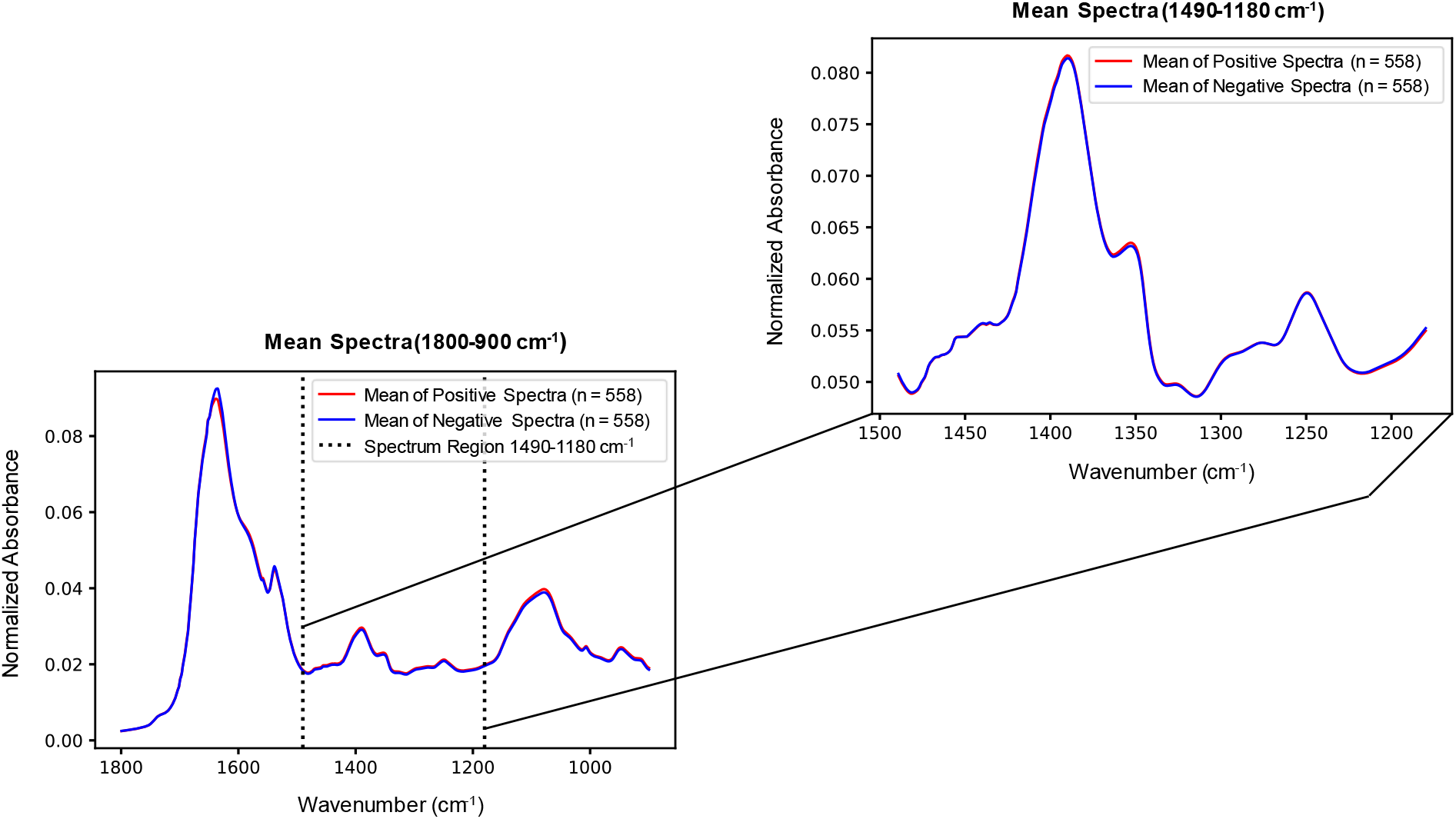
Mean spectra of positive and negative groups from the fingerprint region (1800-900 cm^-1^) and from the spectral region of 1490-1180 cm^-1^. Vector normalization was applied before calculating the mean spectra. Overall, only minor spectral changes were visually observed between the sample groups in the spectral region of 1490-1180 cm^-1^, although the diagnostic performance in the cross-validation setup was almost the same than with the whole fingerprint region (see Figure 2, Table 1, and Table 3 for the spectral region of 1490-1180 cm^-1^ from supplementary material).

## Discussion

The aim of our study was to investigate whether the diagnostic performance of the ATR-FTIR spectroscopy coupled with PLS-DA is adequate to detect SARS-CoV-2 infection from the nasopharyngeal swab samples originally collected and processed for PCR analysis. The AUROC values obtained in our study showed moderate performance (the AUROC values were between 0.66-0.68) when the fingerprint region, and the spectral region of 1490-1180 cm^-1^ were studied. Analyzed dataset contained ATR-FTIR spectra from the 558 negative and 558 positive samples from the separate patients within a real clinical setting in the city of Oulu, Finland.

When analyzing results from our study, it seems that the spectral region of 1490-1180 cm^-1^ is containing enough information to classify ATR-FTIR spectra, even though the whole fingerprint region (1800-900 cm^-1^) is often used in earlier studies. On the other hand, it was not possible to find out separate wavenumbers that could be used to classify spectra. Therefore, at least in this dataset, the meaningful information is spread out over certain spectral region rather than just focused on the separate wavenumbers. Moreover, it is notable that the spectral region of 1490-1180 cm^-1^ was classified with the 13 latent variables, whereas the 31-32 latent variables were needed in case of the whole fingerprint region (1800-900 cm^-1^). Less latent variables mean simpler model that usually refers to better generalization ability. Consequently, in that sense the spectral region of 1490-1180 cm^-1^ could be more suitable choice for spectral analysis.

We observed that only simple spectral preprocessing was needed to prepare the mean spectra for classification, i.e., spectrum truncation to the fingerprint region or the region of 1490-1180 cm^-1^, vector normalization, and spectrum averaging. In practice, this means that our workflow to classify spectra is relatively easy to set up, and the retraining of PLS-DA model in case of more training data available could be also easily accomplished.

Our results with ATR-FTIP spectroscopy showed clearly weaker performance than the results of previous studies with similar methods focusing on SARS-Cov-2 detection. Zhang et al. (10) studied serum with ATR-FTIR spectroscopy using a total of 115 samples (41 of them were confirmed to be COVID-19 positive and others were from healthy donors and patients with other infections or inflammatory diseases). They analyzed the measured dataset with the PLS-DA method and reported the AUROC value as high as 0.9561. Furthermore, coronavirus detection from the saliva (pharyngeal swabs) was conducted by collecting a total of 111 negative and 70 positive samples by combining genetic algorithm with linear discriminant analysis (GA-LDA). Results from that saliva study showed 90% accuracy, 95% sensitivity and 89% specificity (12). Other saliva-based study was performed by asking donors to dribble into a container with added viral transport medium and by analyzing data with the PLS-DA method. From the collected samples, 29 were confirmed as positive and 28 as negative (in overall there were 171 transflection infrared spectra) yielding the sensitivity of 93%, and the specificity of 82% (13). In a very recent study, ATR-FTIR spectroscopy coupled with partial least squares (PLS) and cosine k-nearest neighbours (KNN) analysis was applied to detect SARS-Cov-2 from nasopharyngeal swab (14). There were samples from 243 patients (in total of 714 ATR-FTIR spectra), as where 40 + 111 patients were confirmed as COVID-19 positive. Samples were inserted into the viral transport medium 1 (the liquid 1) or the viral transport medium 2 (the liquid 2). For the liquid 1, the sensitivity was reported to be 84%, the specificity 66% and the accuracy 76.9%. In the case of liquid 2, the sensitivity was 87%, the specificity 64% and the accuracy 78.4%.

There can be various reasons for the overall weaker performance of ATR-FTIR spectroscopy observed in our study. First and foremost, as the nasopharyngeal swab sample was dissolved into a relatively large amount of solvent, the low concentration of viral particles in the viral transport medium (and the viral lysis buffer) may explain the lower diagnostic performance. In addition, it should be taken account that the swab samples were collected also from the people without symptoms but been exposed to COVID-19. It is possible that the performance would be different if the swab samples had been collected from the hospitalized patients of whom swab samples typically contains higher viral loads and the control samples would have been collected from the healthy volunteers.

As a second reason, even though the sample was vortexed before the ATR-FTIR measurements, the sample solution may still have distributed unevenly onto the ATR crystal. This assumption about the uneven viral material distribution may be reinforced by the observation from preliminary test that the averaged spectra (from three repetitive measurements) provided better diagnostic performance when compared to training and validation with each separate spectrum. Consequently, it is possible that the used viral transport medium (and the viral lysis buffer) affects samples causing them to be unevenly distributed, which induces unwanted variation in the ATR-FTIR measurements. In optimal situation, liquid sample drops should be as homogenous as possible to obtain the most accurate results.

It may also well be that the physical sensitivity of ATR-FTIR spectroscopy with these low concentration levels is inadequate to reach the high levels of diagnostic performance. This speculation is also supported by the recent study where excellent diagnostic performance was reported (the accuracy of 90%) when ATR-FTIR spectroscopic measurements were conducted directly from the pharyngeal swab samples, i.e., without dissolving the sample to the viral transport media (12). If that is truly the case, ATR-FTIR spectroscopy could not be recommended as the SARS-CoV-2 screening tool from the nasal swab samples dissolved into the viral transport medium. Instead, the measurement should be conducted directly from the swab stick, or at least without any added chemicals.

Finally, the sample freezing can also affect the performance of ATR-FTIR spectroscopy. It is possible that the freeze-thaw cycle is deleterious for the ultrastructure of viral particles, which would inevitably also affect the sensitivity of ATR-FTIR spectroscopic measurements. Obviously, it would have been the best to conduct all the ATR-FTIR spectroscopy measurements on the same day as the PCR analysis without freezing the samples in between. Unfortunately, it was not logistically possible in this study.

In this study, we used a constant global threshold (0.5) in PLS-DA model to separate between the negative and positive classes. Obviously, if selecting different threshold, the mean accuracies, the mean specificities, the mean sensitivities, and the mean precisions would be different. If portable and fast SARS-CoV-2 detecting test would be developed in the future, the practical choice could be to maximize the specificity, i.e., the ability to detect persons without the disease as effectively as possible. Then persons with negative test result could be diagnosed to be healthy with high confidence. And if positive result would be achieved, one possibility could be then to send a person for PCR test. That would be the case especially if the sensitivity, which describes the ability to detect persons with disease, would be slightly lower.

The indisputable strength of our study is the high number of investigated samples compared to other studies (our study included a total of 1116 separate nasopharyngeal swab samples), including the studies by Nogueira et al. (2021) with a total of 243 nasopharyngeal swab samples in the viral transport medium and Barauna et al. (2021) with a total of 181 undiluted pharyngeal swab samples (12,14). It is well known that the performance values can change, sometimes even drastically, after the preliminary analysis with smaller sample set compared with the eventually larger dataset. Actually, even during our sample collection, we observed better diagnostic performance values when we had collected only less than 200 samples in total (the AUROC values around 0.75-0.78). However, when the sample size increased, the performance values started to stabilize to the current level (the AUROC values between 0.66-0.68). Consequently, more studies with the representative sample sizes (preferably several thousands) are needed to truly validate the ATR-FTIR spectroscopy method for clinical diagnostics of SARS-CoV-2 infection.

## Conclusions

The results from our study with relatively large sample set indicate that the ATR-FTIR spectroscopy coupled with PLS-DA has a potential to detect SARS-CoV-2 infection from the nasopharyngeal swab samples. However, the diagnostic performance of ATR-FTIR spectroscopy remained only moderate, potentially due to low concentration of viral particles in the transport medium (and the viral lysis buffer). Further studies are needed before ATR-FTIR spectroscopy can be recommended for routine fast screening of SARS-CoV-2 from routine nasopharyngeal swab samples.

## Data Availability

Dataset is so recently acquired that it has not yet been published in any repository. All data produced in the present study are available upon reasonable request to the authors

## Supplementary material

## Acknowledgements

Financial support for this research project was received from European Regional Development Fund (ERDF).

## Notes

### Competing Interest Statement

The authors have declared no competing interest.

### Funding Statement

This study was funded by European Regional Development Fund (ERDF) (funding number A76179).

### Author Declarations

The ethical permissions were obtained both locally from the Ethical Committee of North Ostrobothnia's Hospital District as well as nationally from the Finnish Medicines Agency (www.fimea.fi, permission identifier: FIMEA/2021/003461).

